# Detecting respiratory chain deficiency in osteoblasts of older patients

**DOI:** 10.1101/2021.10.06.21264231

**Authors:** Daniel Hipps, Philip F Dobson, Charlotte Warren, David McDonald, Andrew Fuller, Andrew Filby, David Bulmer, Alex Laude, Oliver Russell, David J Deehan, Doug M Turnbull, Conor Lawless

**Author notes:** Corresponding authors &.

## Abstract

Mitochondria contain their own genome which encodes 13 essential mitochondrial proteins and accumulates somatic variants at up to 10 times the rate of the nuclear genome. These mitochondrial genome variants lead to respiratory chain deficiency and cellular dysfunction. Work with the PolgA^mut^/PolgA^mut^ mouse model, which has a high mitochondrial DNA mutation rate, showed enhanced levels of age related osteoporosis in affected mice along with respiratory chain deficiency in osteoblasts. To explore whether respiratory chain deficiency is also seen in human osteoblasts with age, we developed a protocol and analysis framework for imaging mass cytometry (IMC) in bone tissue sections to analyse osteoblasts *in situ*. We have demonstrated significant increases in complex I deficiency with age in human osteoblasts. This work is consistent with findings from the PolgA^mut^/PolgA^mut^ mouse model and suggests that respiratory chain deficiency, as a consequence of the accumulation of age related mitochondrial DNA mutations, may have a significant role to play in the pathogenesis of human age related osteoporosis.

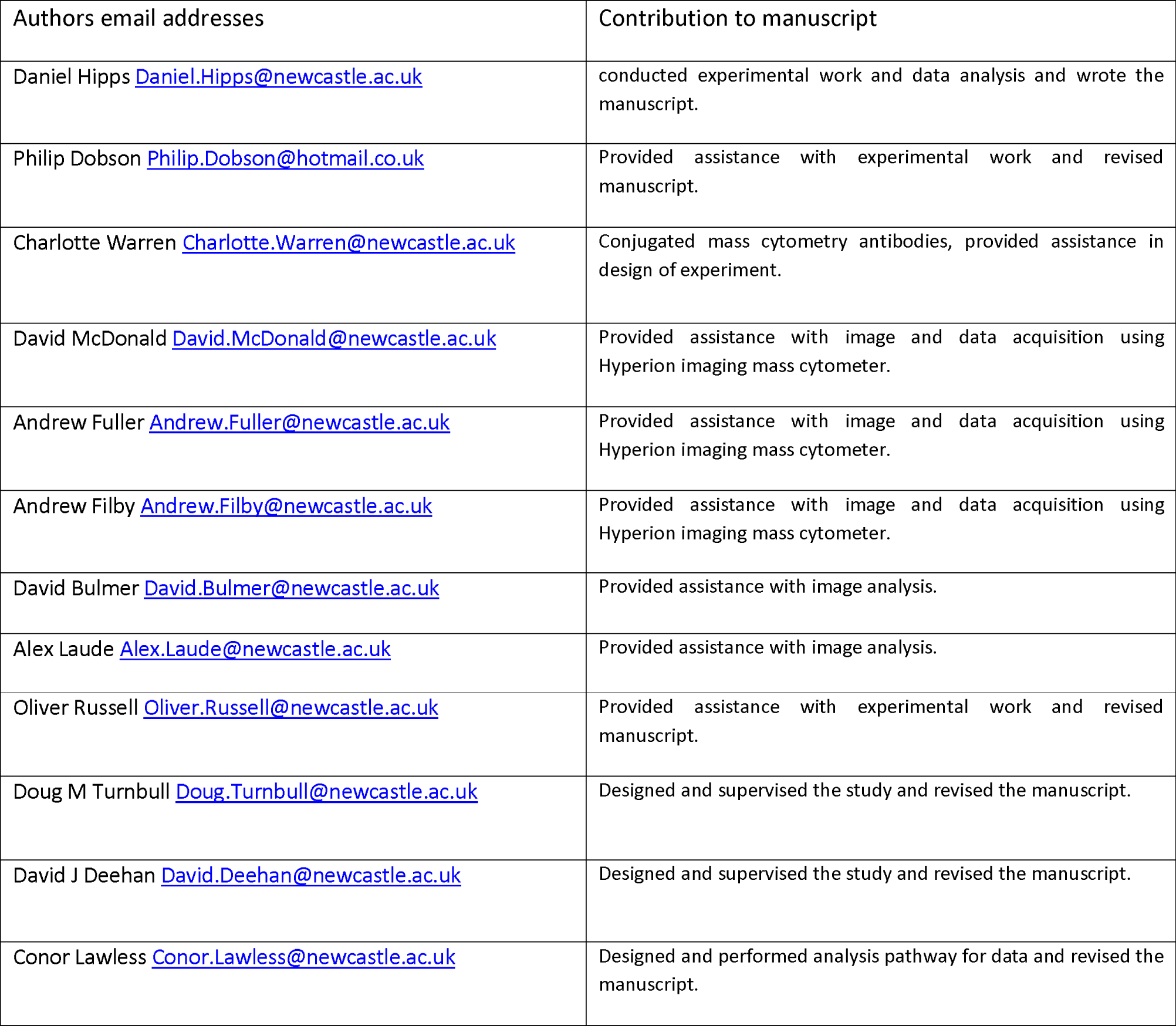

## Introduction

Osteoporosis is a skeletal disease, characterised by reduced bone mass and microarchitecture deterioration, leading to a subsequent loss of strength that predisposes individuals to fragility and risk of fractures with advancing age ^1-3^. Bone mass and mineral density peak in early adulthood, followed by a slow decline as a feature of aging ^4^. This deterioration in bone microarchitecture with increasing porosity and reduced mineralisation levels affects men and women universally. Associated fragility fractures cause significant morbidity and mortality, resulting in significant costs to healthcare economies worldwide ^5-7^.

The pathogenesis of falling bone mineral density and osteoporosis is incompletely understood and is thought to be multifactorial ^4,8,9^. In general, however, age-related osteoporosis is thought to occur because the amount of bone removed by the process of osteoclastic bone resorption is not matched by the amount of new bone formed by osteoblasts ^10-12^. During the production of mineralised matrix, osteoblasts have increased mitochondrial mass in line with the increased metabolic demand of bone mineralisation ^13^.

Humans are known to accumulate mitochondrial DNA (mtDNA) pathogenic variants with age ^14,15^ and these are likely to contribute to the aging phenotype ^16-18^. It has been shown that in the human colon, for example, mitochondrial dysfunction and respiratory chain defects are evident in epithelial crypts from around the age of 30 ^19^. The age at which respiratory chain dysfunction begins coincides with the age at which bone mineral density levels begin to decline in men and women ^4^.

Various mouse models have shown that mitochondrial dysfunction may be integral to the pathogenesis of osteoporosis ^18,20-23^. The PolgA^mut^/PolgA^mut^ mouse model has a defective version of mtDNA polymerase and consequently accumulates mtDNA variants at 3-5 times the rate of wild-type mice and was the first model to demonstrate the causative effect of mtDNA variants in a progeria phenotype ^21^. One of the most prominent features of this phenotype is osteoporosis with a significant reduction in whole-body bone and femur bone mineral density at 40 weeks ^21^.

Subsequent work with the PolgA^mut^/PolgA^mut^ mouse utilising a quadruple immunofluorescent technique demonstrated defects in mitochondrial complex I and IV expression in osteoblasts compared to age matched wild type controls, and also a reduced expression of these components with advancing age in wild type mice ^24^. This mitochondrial dysfunction was shown to be associated with reduced lumbar spine and femoral bone density, reduced bone formation rate, reduced osteoblast population density, increased osteoclast population densities and activity ^25^.

The PolgA^mut^/PolgA^mut^ suggests that mitochondrial dysfunction may have a role in the pathogenesis of osteporosis. Studying human bone using immunofluorescence techniques is more challenging as autofluorescence is relatively higher than in mouse bone. Bone is particularly prone to autofluorescence ^26-29^ due to the concentration of type 1 collagen fibres which luminesce with wavelengths between 400-700nm ^28^. In addition, in more heavily mineralised human bone, autofluorescence is significantly greater ^27^, so signal quality is likely to vary between subjects, depending on individual Bone Mineral Density (BMD). Greater average levels of autofluorescence and greater variability of autofluorescence in human bone make analysis of mitochondrial respiratory chain complex expression within individual cells using fluorescence unreliable and difficult.

Imaging mass cytometry (IMC) offers an alternative method for measuring protein abundance in single cells and is not affected by autofluorescence. IMC relies on the use of stable non-radioactive isotopes of rare earth metals, typically lanthanide metals that are conjugated to antibodies, rather than fluorescent molecules used in traditional immuno-fluorescence detection methods. IMC uses laser ablation of tissue sections combined with time-of-flight Inductively Coupled Plasma Mass Spectrometry (ICP-MS) technology to measure the ions from each cell at a given spatial co-ordinate as the beam raster scans across the selected Regions of Interest (ROIs) in two dimensions. The other major advantage of mass cytometry is that it allows for the simultaneous measurement of a greatly increased number of antibody targets in a single section compared to conventional immunofluorescent techniques ^30^.

To explore whether there is evidence of mitochondrial dysfunction with increasing age in human bone we have developed and optimised an IMC workflow to study mitochondrial proteins in human bone samples. In doing so we can assess the spatial distribution of multiple proteins in single cells within large tissue sections.

## Results

We have developed a workflow, based on IMC, to analyse and assess respiratory chain protein levels within human bone samples at the single osteoblast level. Using metal-labelled antibodies, we were able to target more proteins than is possible in a fluorescent based assay and avoided the severe difficulties with autofluorescence in human bone tissue. With the raw data acquired from the Hyperion module of the Helios mass cytometer we were able to reconstruct pseudo images based upon signal intensity of each antibody (Figure 1). The osteoblasts highlighted in Figure 1D lie along the periosteal border and clear signal can be seen in the equivalent mitochondrial protein channels (Figure 1E-J)

**Figure 1.**
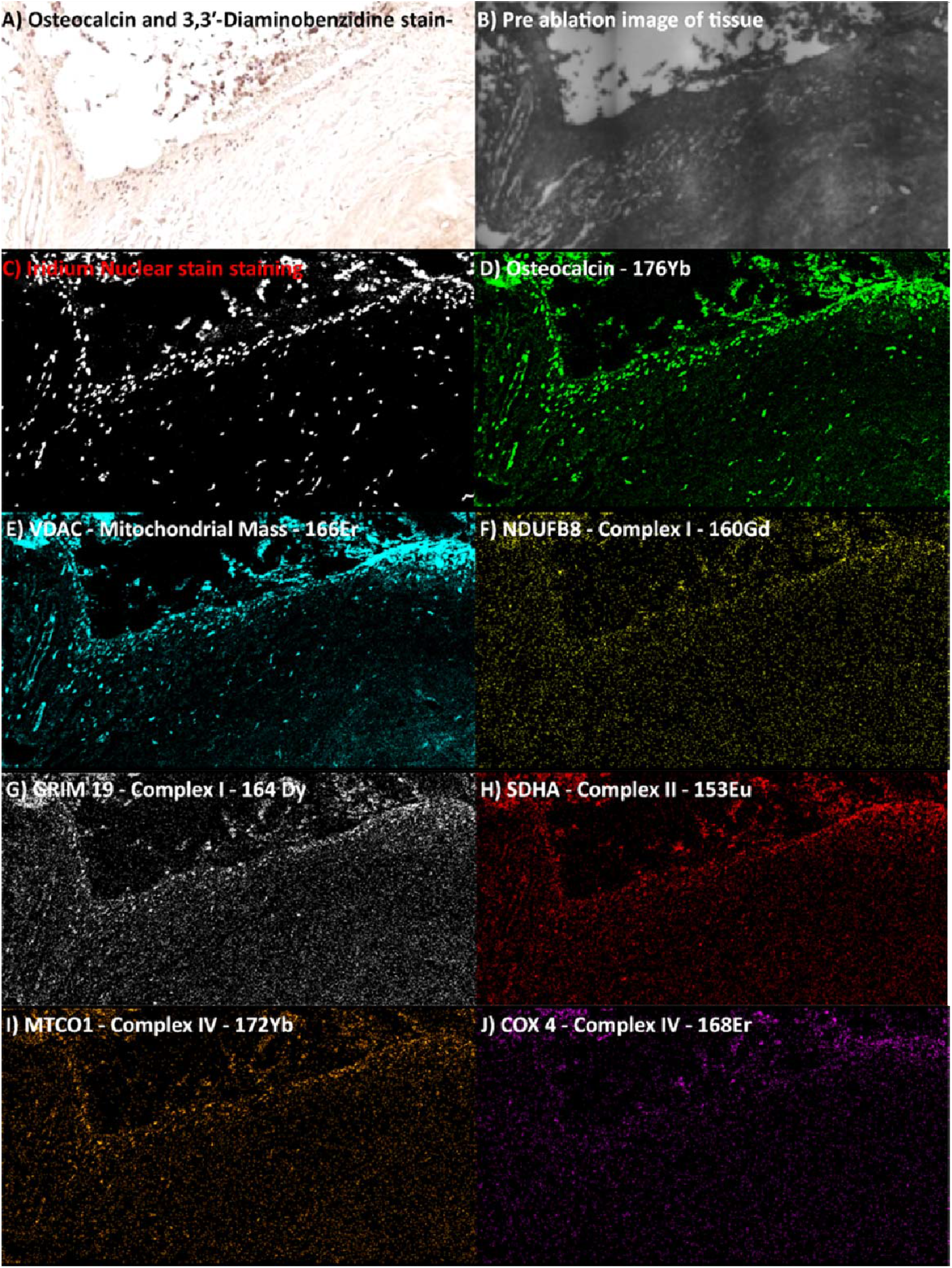
Identifying & measuring regions of interest during IMC. Identifying regions of interest during IMC of bone tissue from patient Hip 10 A) DAB stained osteocalcin positive cells as viewed on Aperio B) – Pre ablation image of the slide as scanned by the Hyperion C) & D) Nuclear staining with Iridium and Osteocalcin, these images were used to identify cells in the analysis steps. E-H) Mitochondrial mass and respiratory chain protein abundance channels from the ablated image after processing with the Hyperion mass cytometer.

**Figure 2.**
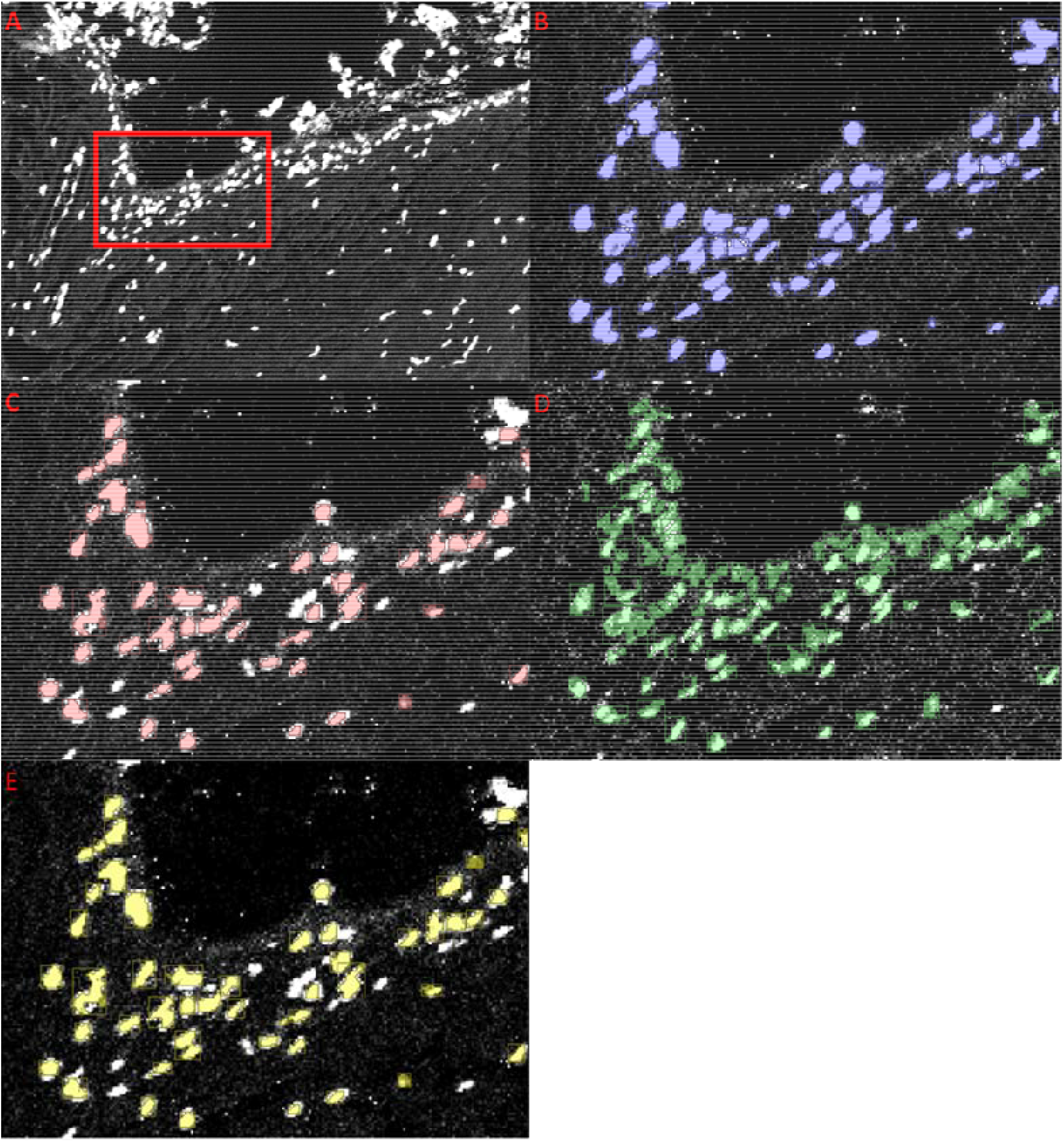
Visualisation of the image segmentation pipeline of ∼75 cells. A) Base image with an enlarged cropped area highlighted in red and in the subsequent images. B) nuclear identification C) Osteocalcin positive pixels associated with a nucleus defining the osteoblast cells and cytoplasm. D) VDAC positive pixels or mitochondrial mass. This image mask is then combined with the mask in panel C E) mitochondrial mass within osteoblasts. Mitochondrial respiratory chain antibody values are then only taken from these positive pixels.

Previous work measuring the expression of respiratory chain complex proteins with mitochondrial mass (VDAC) have been based upon linear regression models accounting for variation in mitochondrial mass and protein deficiency. Unlike this previous work we used 2D kernel density estimates to capture the more complex and diverse shape of osteoblast IMC data.

An example of these kernel density plots can be seen in Figure 3, a male patient aged between 61 & 65 “Hip 3” who showed significant deficiency in complex I proteins (NDUFB8 and GRIM19, 3A and 3B), there was also some deficiency seen in complex II (SDHA, 3C) and ATP synthase (OSCP, 3F). Deficiency in complex IV (COX4 and MTCO1, 3D and 3E) was below 5% and this trend was observed across the data cohort which is summarised in figure 4.

**Figure 3.**
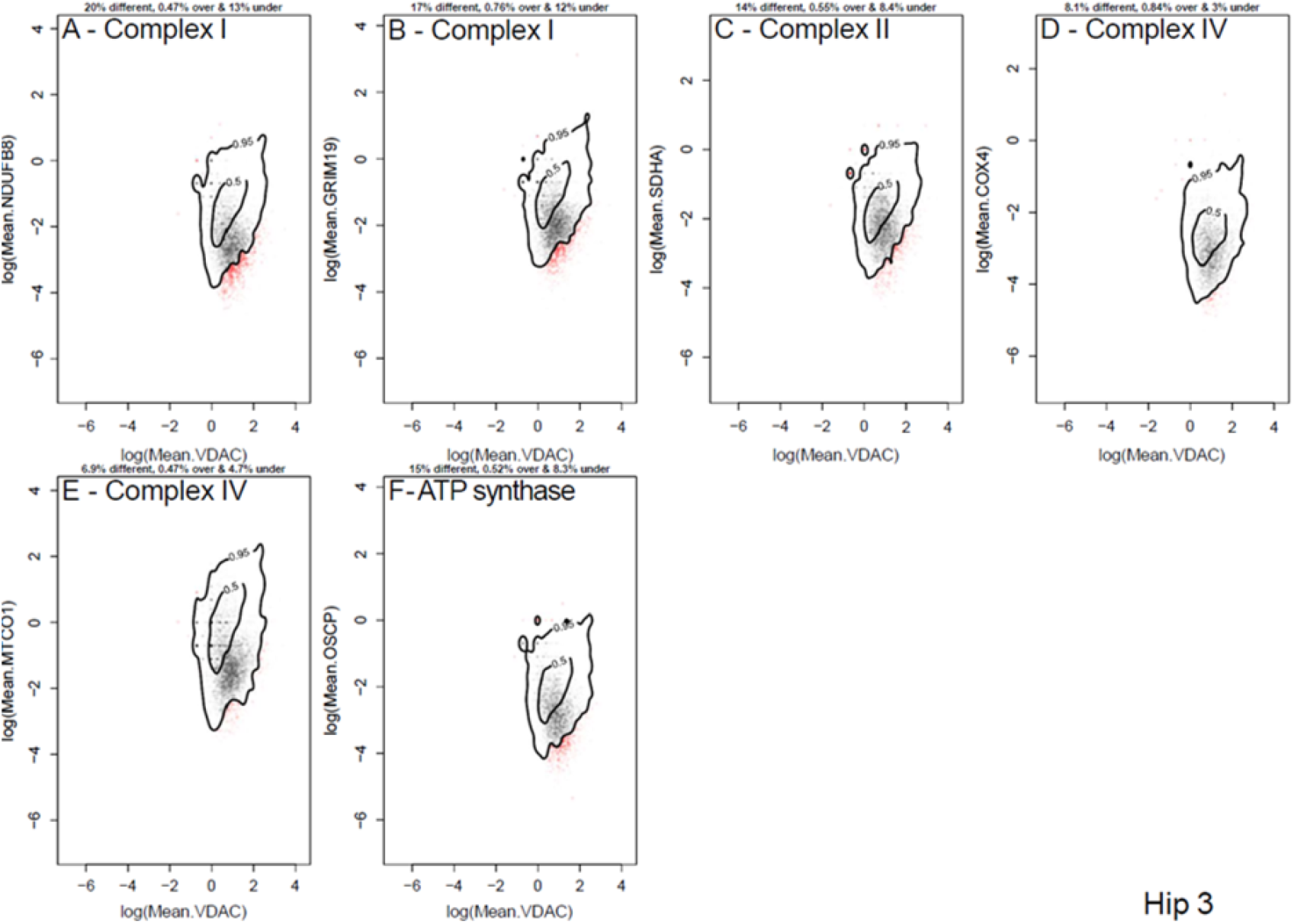
Quantifying RC defects. Data from Hip 3, a male aged between 61 & 65: Contour lines represent the kernel density estimate levels containing 95% and 50% of the control young patient population. Grey points are from patient cells that lie within the 95% contour from controls. Red points are from patient cells lying outside the 95% contour from controls. The proportion of cells different from, vertically above and vertically below the 95% contour from controls written above each panel. A large, significant proportion of cells were deficient in complex I proteins (NDUFB8 and GRIM19, A & B), a significant proportion of cells were deficient in complex II (SDHA, C) and ATP Synthase (OSCP F). The population of cells deficient in complex IV (COX4 and MTCO1, D &E) was below the 5% significance level.

**Figure 4.**
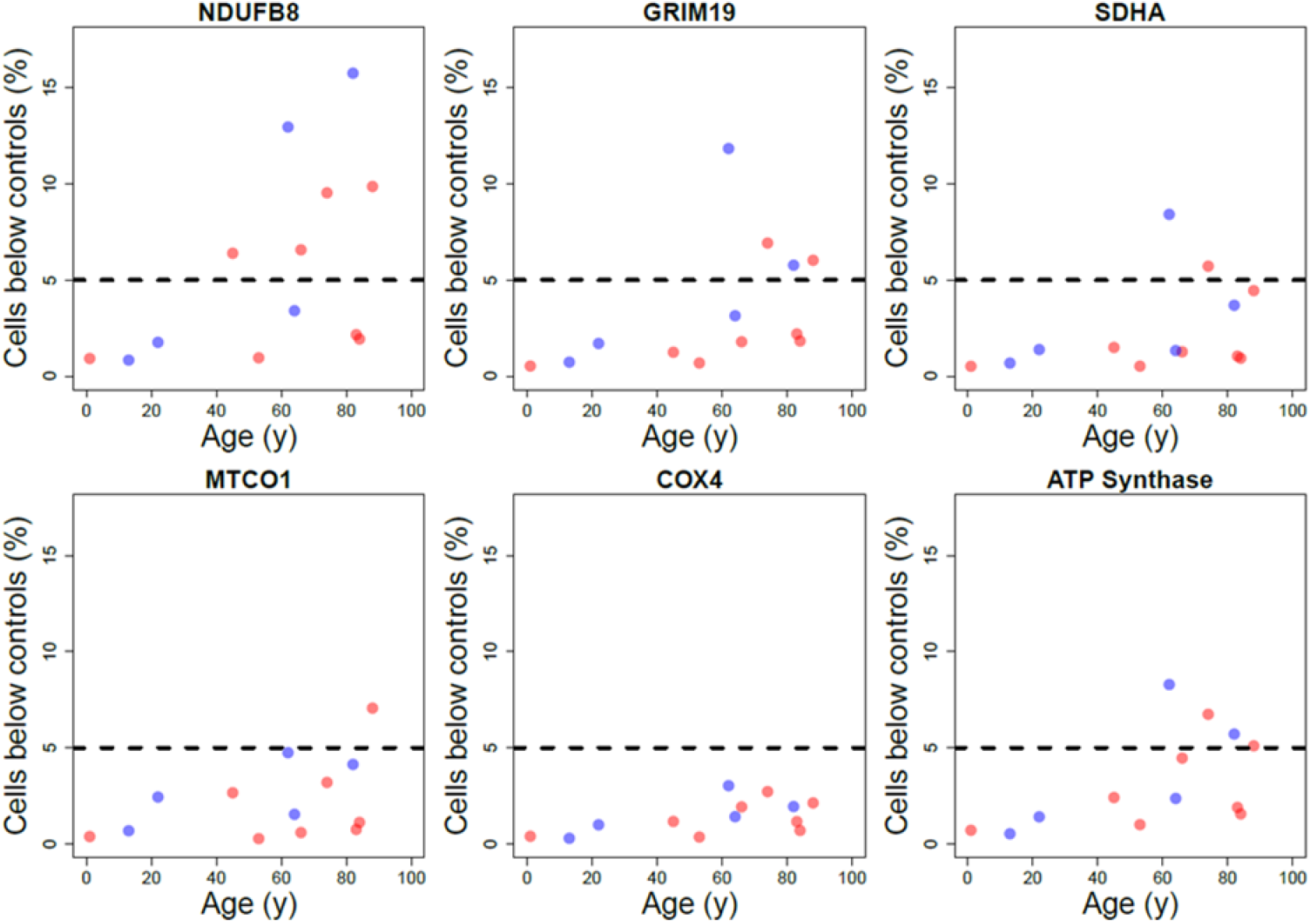
Variation in Complex I (*NDUFB8 and GRIM19*), Complex II (*SDHA*), Complex IV (*MTCO1 and COX4*) and ATP synthase deficiency with age. The proportion of cells lying vertically below the 95% contour from controls in all patients presented in table 2. Blue points represent proportions from male patients and red points from female patients. The horizontal dashed line represents the proportion of differences from healthy control subjects that we might expect to see by chance.

The proportions of osteoblasts with respiratory chain defect increases with age across the full cohort (Figure 4). Specifically, we show an increase in proportion of osteoblasts with NDUFB8 deficiency with age, a less pronounced increase in deficiency with age for an alternative CI subunit GRIM19 and no significant change in expression with age for complex II, IV or V subunits with deficient proportion generally below the 5% cut off for significance across the age range. We found no patient of any age with a significant proportion of fibres above controls (Table 2). The bias towards RC defect and not general deviation above and below control observations demonstrates that the increase in proportion of RC deficient osteoblasts with age is not driven by noise. There is marked variability seen across this small cohort but in general, from around the fifth decade, complex I deficiency becomes apparent. When looking at the individual patients (Figure 4), 5 of the 8 patients over 60 demonstrated significant levels of complex I deficiency whereas only 1 of the 5 patients under 60 had deficiency in complex I. Only 1 patient (a female aged between 86 & 90) had significant deficiency in complex IV. In some patients that exhibited complex I deficiency, higher levels of deficiency were also seen in SDHA and OSCP of complexes II and ATP synthase respectively. There were no patients below the age of 60 that had deficiency in complex II, complex IV or V.

**Table 1.** Antibodies used for imaging mass cytometry

| Antibody | Conjugate | Manufacturer |
| --- | --- | --- |
| Anti-SDHA IgG1 mouse | 153Eu | Abcam |
| Anti-NDUFB8 IgG1 mouse | 160Gd | Abcam |
| Anti-GRIM19 IgG1 | 164Dy | Abcam |
| Anti-VDAC1 IgG2b mouse | 166Er | Abcam |
| Anti-COX4+4L2 Ig2a mouse | 168Er | Abcam |
| Anti-MTCO1 IgG2a goat anti-mouse | 172Yb | Abcam |
| Anti-Osteocalcin IgG rabbit | 176Yb | Santa-Cruz |

**Table 2:** A summary table showing the percentage of osteoblasts above and below the 0.95 contour for complex I, complex II, complex IV and complex V across the patient cohort. Values greater than 5% are considered to be greater than might be expected in control data and are highlighted in red.

| *Patient* | *Age range (years)* | *Sex* | *Osteoblast protein deficiency on CyToF - cells below 0.95 contour* |  |  |  |  | *Osteoblast protein upregulation on CyToF - cells above 0.95 contour* |  |  |  |  |
| --- | --- | --- | --- | --- | --- | --- | --- | --- | --- | --- | --- | --- |
|  |  |  | NDUFB8 | GRIM19 | SDHA | MTCO1 | COX4 | NDUFB8 | GRIM19 | SDHA | MTCO1 | COX4 |
| ***Paediatric 1*** | 1-5 | F | 0.92% | 0.53% | 0.52% | 0.38% | 0.39% | 2.00% | 3.30% | 3.30% | 2.20% | 2.80% |
| ***Paediatric 2*** | 11-15 | M | 0.84% | 0.73% | 0.68% | 0.68% | 0.29% | 0.76% | 1.50% | 1.10% | 0.97% | 0.86% |
| ***Paediatric 3*** | 16-20 | M | 1.80% | 1.70% | 1.40% | 2.40% | 0.99% | 1.10% | 1.20% | 1.40% | 0.67% | 0.97% |
| ***Hip 1*** | 41-45 | F | 6.40% | 1.20% | 1.50% | 2.70% | 1.20% | 0.46% | 0.91% | 1.80% | 0.54% | 2.90% |
| ***Hip 2*** | 51-55 | F | 0.96% | 0.69% | 0.52% | 0.27% | 0.35% | 2.80% | 4.10% | 4.80% | 2.50% | 3.60% |
| ***Hip 3*** | 61-65 | M | 13.00% | 12.00% | 8.40% | 4.70% | 3.00% | 0.47% | 0.76% | 0.55% | 0.47% | 0.84% |
| ***Hip 4*** | 61-65 | M | 3.40% | 3.10% | 1.30% | 1.50% | 1.40% | 1.60% | 1.40% | 1.90% | 0.46% | 1.10% |
| ***Hip 5*** | 66-70 | F | 6.60% | 4.50% | 1.30% | 0.59% | 1.90% | 0.52% | 1.20% | 1.10% | 0.68% | 1.30% |
| ***Hip 6*** | 71-75 | F | 9.50% | 6.90% | 5.70% | 3.20% | 2.70% | 2.20% | 4.20% | 2.20% | 2.90% | 2.70% |
| ***Hip 7*** | 81-85 | F | 16.00% | 5.80% | 3.70% | 4.10% | 1.90% | 1.10% | 0.97% | 1.40% | 0.32% | 1.40% |
| ***Hip 8*** | 81-85 | F | 2.20% | 1.90% | 1.00% | 0.75% | 1.20% | 0.88% | 1.10% | 1.30% | 0.67% | 0.78% |
| ***Hip 9*** | 81-85 | F | 1.90% | 1.80% | 0.95% | 1.10% | 0.69% | 0.67% | 1.20% | 1.00% | 0.72% | 0.68% |
| ***Hip 10*** | 86-90 | F | 9.90% | 5.10% | 4.50% | 7.10% | 2.10% | 1.00% | 1.40% | 1.10% | 0.70% | 1.10% |

## Discussion

We have demonstrated the application of IMC to measure respiratory chain protein expression within bone in individual osteoblasts. We were able to analyse over 130,000 cells across samples from 13 patients. Six proteins and two DNA markers were detected using our antibody panel but IMC has the scope for many more antibodies to be detected synchronously on the same slide. This forgoes the need for further multiplexed staining methods requiring further slide preparation or sequential staining steps which can affect the integrity of the tissue. Along with antibody data, IMC also has the scope to provide spatial data for the selected antibodies with the co-ordinates of each ablation location recorded along with the signal and reconstructed into an image. Analysis of fixed FFPE and snap frozen tissue allows flexibility and the retrospective analysis of archived samples ^31,32^.

The staining protocol is relatively simple and based upon an established quadruple immunofluorescent assay ^24^. This translation from immunofluorescence to IMC is relatively straightforward and could be replicated for other protein targets or tissues. We found that conventional tools such as CellProfiler ^33,34^, was not suitable for IMC of osteoblasts due to the heterogeneous nature of our samples and relatively small pixel areas. Using image reconstructions or TIFFs created from the raw data using Fluidigms MCD viewer, we were unable to reliably segment individual osteoblasts to allow single cell analysis using CellProfiler, or with mitocyto ^35^ which was developed for analysis of IMC images of skeletal muscle fibres.

We have developed a unique image analysis pipeline that permits analysis of a very large number of osteoblasts (≈160,000 in this work). However, the data was found to be considerably noisier than IMC or quadruple immunofluorescence data from much bigger and more clearly defined skeletal muscle fibre sections. This is due to the small cell size and relatively lower cytosol to nuclear size ratio, with each pixel representing a greater proportion of the cell than in larger cell types. IMC data is also nosier than light microscopy as the resolution is lower and acquisition times are linked to laser output. Using a decreased laser ablation spots per second would improve spatial resolution but reduce the sensitivity of acquisition as less ions would be collected but the time taken for acquisition would also be longer. In contrast having more laser ablations spots per second would increase the speed of acquisition but also the risk of contamination between spots ^36^. IMC requires a trade-off of these properties to get accurate data in an appropriate time frame. It is this noise and the size of the cells we were analysing which made segregation of the cells so important and why a new method to analyse this novel technique had to be developed.

The method of analysis developed here is a clear pathway that can be used in heterogeneous tissue with some degree of automation once the parameters to determine cell and protein detection are appropriately defined. Most IMC data generated thus far has been on relatively homogenous cellular soft tissue such as tumours and the immune system ^32,37^. Here IMC has been utilised to discover the link between cell biology and disease ^31^. The significant advantage of imaging mass cytometry over more conventional techniques, and even mass cytometry, is that it allows the simultaneous measurement of multiple features in multiple cells, all within their microenvironment without disruption of tissue architecture.

The protocol and image analysis pathway presented here has considerable potential for other investigations using human bone. With none of the problems associated with autofluorescence, the ability to analyse many more proteins and individual cells without disturbing the microarchitecture of mineralised bone is now possible. Whilst IMC represents an important advance, there are limitations. To date most conjugated antibodies available are targeted towards cancer and immunology antigens. To address this specific protein free antibodies need to be used and these antibodies can be conjugated to heavy metal tags using a conjugation kit from Fluidigm ^38^.

The major drawback of IMC is the time to ablate tissue: around 0.75mm^2^ per hour. Due to the amount of time taken to image sections, target areas or ROIs were used rather than imaging the whole slide. In comparison to soft tissues, the solid-state laser settings needed to be at their maximum intensity to ensure adequate ablation of bone tissue. The other limiting factor is the resolution which is approximately 1μm^2^/pixel. Whilst on the whole, this allows subcellular imaging, the resolution is poor in comparison to microscopy and visualising subcellular detail such as mitochondrial shape and morphology is not possible, especially in smaller cells like osteoblasts (20-30 μm diameter). In smaller cells this resolution becomes more problematic: a greater proportion of the cell is covered by a single pixel. A combination of signal and background in single pixels could lead to under representation of the antibody target as the pixels do not create a continuous border along the cell margin.

Whilst autofluorescence is not an issue using this protocol, background signal is important to consider. In terms of specificity of the signal, as in fluorescent based techniques, non-specific binding of antibodies can be an issue. For this reason, we based the protocol upon validated and tested techniques using antibodies with good specificity employed in fluorescent based analysis prior to conjugation with heavy metals.

Our approach was to carry out noise reduction steps as part of the Volocity analysis pipeline. Signal was also only recorded for areas identified by the various masks created through Volocity. Other authors have taken their own individual steps to address the issues of background. A standardised method is currently not clear and is image and tissue dependent. One group have utilised a 5 × 5 μm^2^ median filter to account for horizontal streaking: for each pixel, if its intensity value is in the top 2% of pixels it was removed ^39^. Whereas another group used a “no primary antibody” type control step measuring the background signal of a channel not associated with antibody signal in their IMC work. The signal intensity was then used as a threshold to remove background signal intensity. Areas of isolated low signal intensity (which were filtered out) were associated with noise whereas true signal was clustered ^40^.

Using this assay, we aimed to show a link with age and accumulated mitochondrial respiratory chain protein deficiency in osteoblasts. The antibody targets chosen were those of the mitochondrial respiratory chain subunits that have been well validated from previous studies ^24,35,41^. The patients were across a spectrum of ages (0-90 years). This sample population and distribution between the extremes showed an increase in mitochondrial respiratory chain complex I deficiency with age. This is in line with previous research in other human tissues ^15,17-19,42^ and the POLG mouse model ^24^.

Mitochondrial DNA is prone to mutations with age with *de novo* variants arising in mtDNA at a rate up to 10 times greater rate than in nuclear DNA ^43^. Complex I is the most frequently observed mitochondrial respiratory chain complex to be deficient and this in is in part due to its structure. It is the most complicated of the complexes in terms of its components and assembly. It is coded for both by nuclear and mitochondrial DNA, consisting of 45 subunits of which 7 are encoded by mtDNA ^44^. All 7 of these mtDNA coded proteins have reported pathological variants as do 21 of the nuclear encoded proteins and 10 of the assembly factors ^45,46^. The mtDNA coding for complex 1 is many more base pairs than the other mtDNA encoded proteins of the other complexes and so pathogenic variants are more frequent.

Although our data shows significant differences in the complex I protein, NDUFB8 between young and old patients, the effect is not so apparent in the limited sample population for GRIM19 (NDUFA13). GRIM19 is part of the P module of complex I and is among the first proteins assembled as part of the Q/Pp-a intermediate and only contains 1 mtDNA encoded protein (ND1). NDUFB8 is part of the PD-b module which is made up of the mitochondrial encoded proteins ND2, ND3, ND4-L, ND5 and ND6 ^47^. Thus, issues affecting the assembly and incorporation of NDUFB8 into complex I have a greater chance of causing deficiency than those of GRIM19 due to the presence of more mtDNA encoded proteins in each individual intermediate’s assembly.

Changes in complex IV in previous studies are not detectable below the age of 35 and only detectable in one-third of osteoblasts in patients over 70 ^14^. We did not detect significant levels of complex IV deficiency in the osteoblasts of our aged patients. Complex IV tends to be less affected by sporadic mtDNA mutations, possibly due to the lower number of Complex IV genes encoded by mtDNA. Complex IV has just three mtDNA coded proteins (COI-III). This accounts for 27% of the Complex IV proteins compared to 56% of Complex I proteins ^48^. Not only is mtDNA more heavily involved in coding for Complex I proteins but age-related mtDNA mutations frequently affect the area between the heavy and light strand origins of replication ^49^. This frequently affects the ND3, ND4, ND4L and ND5 proteins which are components of complex I. Overall pathogenic variants affecting complex I are around twice as likely to occur than those affecting complex IV ^48^.

This work highlights the correlation between accumulating mitochondrial respiratory chain deficiency and advancing age in human osteoblasts. The PolgA^mut^/PolgA^mut^ mouse model showed evidence of premature osteoporosis and associated osteoblast respiratory chain deficiency was confirmed previously and shown to be associated with declining bone density ^24,25^. *In vivo* assays demonstrated significant defects in bone formation by ostoeblasts with reduced osteoblast numbers, increased osteoclast numbers, and *in vitro* assays demonstrated increased resorption activity by osteoclasts.

Osteoporosis pathogenesis is multifactorial. The new discovery in terms of quantifying the increasing presence of respiratory chain deficiency in human osteoblasts offers an insight into the pathology and cause of age-related osteoporosis at a protein-based and cellular level. This mirrors the findings of the PolgA^mut^/PolgA^mut^ mouse model and offers an insight into the pathogenesis of osteoporosis where increased bone resorption and decreased bone formation occurs in conjunction with mitochondrial dysfunction. This relates to age acquired osteoporosis and how mitochondrial respiratory deficiency may be integral to its development, in keeping with the mitochondrial theory of ageing which affects cells almost universally ^43^.

Current treatment regimes are targeted and modifying risk factors in terms of vitamin D and calcium deficiency and the use of bisphosphonates which prevent osteoclastic resoprtion but do have significant side effects and longterm risks ^50^. Understanding the pathology of osteoporosis at a protein based metabolic level paves the way for further research and subsequent treatment developments. We have pioneered a method which allows accurate quantification of respiratory chain activity within individual osteoblasts *ex vivo*, further in depth investigation of the role of mitochondrial function on various bone pathology.

## Conclusion

We have optimised and developed the first use of an IMC assay on human bone assessing multiple proteins in osteoblasts. We have demonstrated the suitability of IMC to analyse highly heterogeneous tissue and bone across a range of ages, assessing increased respiratory chain deficiencies present in human osteoblasts with age. This has further potential to build upon previous work linking mitochondrial respiratory chain activity with the pathogenesis of osteoporosis ^24,25^.

Further development of the use of IMC in bone has the potential to enable multiple synchronous assays, in contrast to previously limited techniques using limited antibodies, and also avoids constraints such as issues caused by fluorescent based analysis in bone.

## Materials and Methods

### Sample collection

Samples were collected from patients undergoing routine elective orthopaedic surgery from the Newcastle upon Tyne Hospitals. Ethical approval for the collection of paediatric samples was via the Great North Biobank, application number (*GNB-012*). Ethical approval for use of adult human samples was gained as an adjunct to the Newcastle Bone and Joint Biobank – (*REC reference 14/NE/1212, IRAS project ID 166522*). Newcastle University reference 8741/2016.

Samples of bone were taken from the femoral neck at the time of total hip surgery. Following removal of the femoral head with an oscillating saw, a wafer of femoral neck, approximately 5mm thick was collected using an oscillating saw. To avoid cells which may have been subjected to thermal damage during this process, subsequent tissue samples were taken from the central portion of this wafer.

Paediatric samples were obtained at the time of pelvic osteotomy for developmental dysplasia of the hip (Paediatric 1) and when “bony bridges” were excised following a physeal injury and growth arrest (Paediatric 2). Both of these samples were taken with osteotomes (chisels) so risk of thermal necrosis was avoided. A further younger patient sample was also taken at the time of femoral intramedullary nail insertion in a patient aged between 16 & 20 (paediatric 3)

### Fixation and Decalcification protocol

Samples were trimmed and placed in tissue cassettes, fixed in 4% PFA for 72 hours followed by a dH_2_O wash. Decalcification was performed using 14% tetra EDTA buffered to pH 7.4 at 4°C with agitation. The solution was changed three times per week for 21 days. Following this, samples were washed with dH_2_O and embedded in paraffin. 4μm sections were cut on to Leica X-tra slides, left to oven dry for 48 hours and then air dried for one week. In our experience these drying steps minimised tissue loss in the subsequent staining steps.

### Maxpar and imaging mass cytometry antibodies

The antibodies used for this study are listed in table 1. The mitochondrial antibodies were optimised in previous work from our laboratory and their signal strengths have been shown to correlate with functional changes ^35^. Antibodies were paired with heavy metals for conjugation based on fluorescence intensity in immunofluorescent labelling with a 488-secondary antibody. Antibodies with a higher signal on immunofluorescence were paired to metals known to give a weaker signal on IMC and *vice versa* as per previous published work ^35^. Anti-Osteocalcin IgG rabbit was conjugated to Yb-176 using the Fluidigm antibody conjugation. The efficacy of this antibody was confirmed with DAB staining. Successful conjugation was also confirmed using antibody capture beads (AbC™ Total Antibody Compensation Bead Kit A10497) on the Helios mass cytometer. Intercalator-Ir (Fluidigm 201192A) is a cationic nucleic acid intercalator. It contains a natural abundance of the iridium isotopes 191Ir and 193Ir and was used as a nuclear marker at 0.3125μM.

### Imaging mass cytometry staining protocol

Serial sections were cut from paraffin blocks. Antibodies to osteocalcin were applied to the first section which was stained with 3,3⍰-Diaminobenzidine (DAB) and scanned with the Leica Aperio imaging system to use as a reference when identifying sites to ablate.

The serial sections for IMC were first dewaxed for 30 minutes at 60°C. Residual paraffin was then cleared with two five-minute clearing steps in Histoclear before graded rehydration through an alcohol gradient. Antigen retrieval was performed as described previously ^51^ using 1mM EDTA at pH 8.0 at 80°C for 35 minutes ^24^. Sections were then rinsed in cold _di_H_2_O (deionized water). Blocking was performed with a 10% NGS in PBS for one hour at room temperature. Sections were then once again rinsed in cold_di_H_2_O before applying antibodies.

All conjugated antibodies were mixed to create a cocktail before dilution at 1:50 in 10% NGS and PBS. The antibody cocktail was placed on sections which were then incubated in a humidified chamber at 4°C for 12 hours. Following PBS wash, iridium-intercalator was used at 0.3125μM in PBS for 30 minutes at room temperature to stain the nuclear regions. This was followed by a further wash step in _di_H_2_O for 7 minutes. Slides were then air-dried at room temperature for 20 minutes before being stored, ready for ablation.

The slides were then loaded into the Hyperion imaging module of the Helios mass cytometer (Fluidigm). A provisional brightfield image was taken and loaded into the software (Figure F1B). From this image and by comparison with the DAB stained image (Figure 1A) on Aperio, “panoramas” were created containing areas with osteoblasts. Within the panoramas, ROIs were selected based on expert pathology input and subsequently ablated. These panoramas were then scanned with the Hyperion and the ROIs were ablated to give pseudo-images representing expression of each channel (Figure 1 C-J).

### Analysis Pathway

We used MCD viewer to view images and perform a qualitative QC of each stain. We then exported images as 16-bit TIFFs with each channel from each ROI exported as a separate TIFF file. The TIFFs were combined in Nikon Elements, converting the multi-channel TIFFs into a single multi-level ND2 Nikon Elements file that could then be imported into the Perkin Elmer software Volocity 6.0 for analysis. Raw, multi-channel .tiff files can be accessed here: https://doi.org/10.25405/data.ncl.16686952

To quantify protein expression in each cell, the multi-channel images were cropped to the areas of interest within each ROI. The cells were then identified using the nuclear marker (Ir-191/DNA 1, Figure 2B), an exclusion based on size and a split of touching objects was then applied. The cell cytoplasm was then identified by using the osteocalcin cell marker (channel 176, Figure 2C). A mask was then created to highlight each cell by combining the osteocalcin signal associated with a nucleus identified in the first step. Within this area, a further mask was created by identifying only the VDAC positive pixels indicating areas of mitochondrial mass (Figure 2E). Masks were then used to quantify protein expression in individual cells. A noise reduction filter was also applied to limit background signal from IMC noise. Overview images of this process can be seen in Figure 2.

To estimate the proportion of cells deficient in one or more proteins, we adapted an established muscle fibre workflow ^41^. We used a non-parametric 2D kernel estimate of the density of cells from healthy patients in NDUFB8-VDAC space, generated using the kde2d function from the MASS package in R ^52,53^. Specifically, we calculated the contour level from the KDE which contains 95% of the control cells. Then, for each patient, we annotate any cell which lies outside that 95% contour as being “different” from controls. We defined control cells as those acquired from the ablation of samples from paediatric 1, 2 and 3. We further label cells lying “above” (e.g. with NDUFB8 values vertically above the contour) or “below” (e.g. with NDUFB8 values vertically below the contour). Code and data for calculating proportion of cells classified as normal, “above” and “below”: https://github.com/CnrLwlss/Hipps_IMC.

## Data Availability

Raw image data is hosted permandently at data.ncl.ac.uk
Tabular summary data (& code for analysis) is hosted permanently on github.

https://doi.org/10.25405/data.ncl.16686952

https://github.com/CnrLwlss/Hipps_IMC

## Acknowledgements

This work was supported by The Wellcome Centre for Mitochondrial Research, Newcastle University Centre for Ageing and Vitality (supported by the Biotechnology and Biological Sciences Research Council and MRC [G016354/1]), the Malhotra Group, Newcastle NIHR Biomedical Research Centre in Age and Age Related Diseases award to the Newcastle upon Tyne Hospitals NHS Foundation Trust. The Royal College of Surgeons, England provided a one year Fellowship to support this work.

## Contributions

D.H conducted experimental work and data analysis and wrote the manuscript. P.F.D and O.R provided assistance with experimental work and revised manuscript. C.W conjugated mass cytometry antibodies, provided assistance in design of experiment. D.M, A.Fu and A.Fi Provided assistance with image and data acquisition using Hyperion imaging mass cytometer. D.B and A.L provided assistance with image analysis. D.M.T and D.J.D Designed and supervised the study and revised the manuscript. C.L designed and performed analysis pathway for data and revised the manuscript. All authors approved the final submitted version.

